# ‘You are too much in this modern world, that’s why you are like this’: Understanding perceptions of mental health among Somali people in London

**DOI:** 10.1101/2024.02.28.24303492

**Authors:** Caitlin Gonzalez, Elizabeth Humberstone, Chris Willott

## Abstract

This is a qualitative study exploring the perceptions of mental health among Somali people living in London. Participants, over the age of 18 and identifying as Somali, were recruited from a community centre in West London. Seven participants were recruited, and semi-structured interviews were conducted to better understand perceptions of mental health, care-seeking, treatment and different understandings of these issues within their community. Interview transcripts were imported into NVIVO version 14 to be coded, and description-focussed thematic analysis was used to interpret key themes.

Concerns around judgement, shame and stigma are key issues affecting attitudes towards mental health in this group. Other important issues affecting conceptualisations and attitudes towards mental health were intergenerational differences, isolation from the community, stigma and secrecy. Participants also reported the importance of protective factors, particularly faith and family in their lives. This research adds to existing literature in exploring perceptions of mental health in Somali communities in the UK, and such research is helpful in identifying cultural barriers to recognition and treatment of mental health within this community.

## Introduction

### Background and Context

According to the 2021 United Kingdom (UK) census, there are approximately 176,000 people living in England and Wales who identify as Somali [1]. Greater Somalia has experienced civil unrest since the start of the war in 1991, with millions of Somalis fleeing to parts of Africa, Europe and North America. Individuals fleeing conflict are significantly more likely to experience mental health problems, due to trauma, settlement stress and social isolation and higher prevalence of anxiety and depression is reported in Somali people in comparison to the general population in Western countries [2,3]. Furthermore, research shows that mental health conditions are heavily stigmatised within Somali communities in the West, including in Norway [2], the United States [4] and the UK [5].

Somali people trace their origins to the Horn of Africa, and the main Somali population in Africa stretches across four countries (Somalia, Ethiopia, Kenya and Djibouti) and one *de facto* independent state (Somaliland). The region has been the site of numerous conflicts since the early 1980s, initially motivated by resistance to the military junta in the Somali Democratic Republic led by Mohammed Siad Barre [6]. Since then, numerous international, national and subnational conflicts involving state and non-state actors have affected the lives of millions of people. Conflict in the region continues to this day, alongside significant climate change-induced flooding [7]. Conflict and environmental change have caused mass emigration from the region, with displaced Somali people seeking refuge in other countries; whilst most have claimed asylum in neighbouring African countries, there are also large numbers of Somali refugees in Europe [8,9].

**Figure 1:**
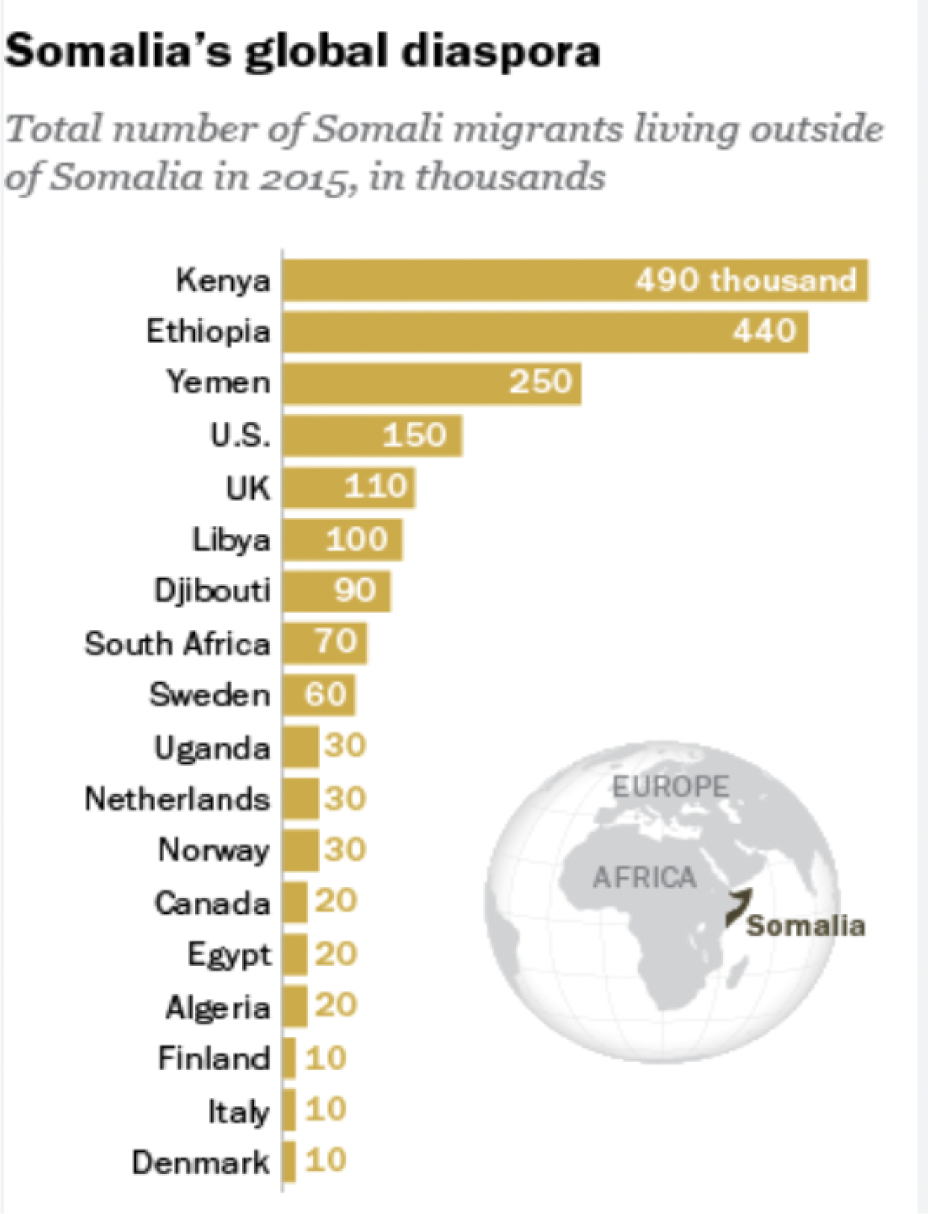
Somali diaspora (Source: Pew Research Centre, 2016)

### Somali diaspora in the UK and their relationship to mental health

Although official counts refer only to Somali people from the Federal Republic of Somalia, rather than ethnic Somalis from other African countries, evidence suggests that the UK is home to the highest number of Somali people in Europe [5].

Research conducted in the UK has found a high proportion of Somali people having unmet needs in key areas, such as housing, food, physical health, psychological distress, and educational attainment [10]. Through discrimination and adversity, these factors, alongside experiences of conflict, make Somali people more susceptible to mental health symptoms [11,12].

Understanding Somali people’s perceptions of mental health is crucial to creating culturally sensitive mental healthcare services, an area where their needs are currently unmet [11,13,14]. Mental health service use has been noted to be low in Somali populations living in London, despite a high proportion of the community reporting mental health problems [10]. Mental health service use for Somali people in Western countries may be limited due to a combination of stigma, lack of awareness at the community and family level and cultural preferences in seeking help in religion and community [15]. Thus, research into Somali attitudes toward mental health that explore the barriers to seeking formal mental health services, and seek opinions on helpful routes for treating mental health could help support and develop services to become more culturally appropriate. Amongst the Somali diaspora, cultural, religious and moral values contribute to a strong sense of identity [16]. Religious practices, peer opinion and family support may all impact perceptions of mental health, and it is generally expected that individuals would seek support from the community before engaging with formal mental health services [16,17].

Within Somali vocabulary there is an absence of words for psychological diagnoses such as depression or stress. This lack of translation, alongside a lack of knowledge on Western psychological concepts, create difficulty around conversations concerning mental health [16]. Furthermore, research has suggested that restriction in vocabulary may contribute to a practice where individuals describe somatic symptoms such as headaches, tiredness and heart palpitations to express their mental distress [18]. Understanding such customs could therefore help support clinicians in primary care services with assessing and managing Somali patients appropriately.

Due to unfamiliarity with Western psychiatric labels, mental health is frequently viewed as binary; an individual is either ‘sane’ or ‘insane’ with no grey area in between, and those with mental health conditions frequently labelled ‘mad’ or ‘crazy’ [5,19]. This dichotomy could contribute to stigma surrounding mental health, a concern that is noted within the community [17]. People with mental health conditions are also frequently seen as a risk to the community, further exacerbating stigmatising attitudes [11]. Consequently, individuals may be ostracised for speaking about their mental health condition or treatment, further reinforcing a cycle of shame, stigma and isolation [17,19].

## Materials and methods

### Study setting and rationale

In this article, we analysed the perceptions of mental health among Somali people living in London. The study participants all attend Ilays Community Centre in Feltham or are connected to the centre in some way. Ilays is a non-profit organisation that helps BAME communities arriving in the UK and was set up in 2004 by the East African community in West London [20].

Most current research on Somali communities in the Western world focuses on the United States. This research has identified a gap in the literature by further exploring perceptions of mental health among Somalis living in London. It will aid the understanding of the changes necessary to improve mental health services for this population group, and will contribute to knowledge of the perceptions, beliefs and practices prevalent within the Somali community, while recognising culturally specific perspectives that affect help-seeking behaviours.

### Research paradigm and question

The main goal of the article is to analyse the meanings people attach to mental health based on their experiences. A qualitative approach was chosen as this provided an unconstrainted, flexible and open environment for participants to respond to questions considering the nature and subject of the research [21].

This research uses an interpretivist approach where phenomena are observed through subjective accounts, perceptions and opinions based on the individual’s perception of the world in different contexts, allowing the researcher to create meaning and synthesise perceptions of mental health [22,23]. The ontological approach assumes multiple truths and realities and suggests that perceptions of mental health are socially constructed and subject to constant change [22]. This research is therefore intended as a sample of the views of Somali people living in London, where the opinions and views expressed in this article are developing and changing.

### Data collection and sampling

Purposive sampling was used to recruit participants who are over 18 of Somali ethnicity and living in London. This sampling method was used as it aims to select respondents who are most likely to provide useful and appropriate information in interview [24]. Ilays was chosen as the basis for recruitment as this is a hub for Somali community activities in West London. CG visited the community centre several times to recruit participants, using a poster. This was not successful so staff at the Centre put CG in touch with potential participants who were followed up with an information sheet.

This study recruited seven participants, who were all women. All participants were under the age of 60, and all but one had migrated to London from Somalia, either at a young age or as parents with their children. Semi-structured interviews with these participants were conducted. Six interviews were conducted over the phone, which offered flexibility and ease for participants given that most participants were mothers and lived in different areas in London [25]. One face-to-face interview took place at the Ilays community centre. Data was collected in June and July 2023. All interviews were carried out by CG.

### Data analysis

Collected data was imported into NVIVO version 14 to be coded. Description-focused coding was used. Thematic analysis was used to interpret the data by identifying themes and patterns based on common perceptions of mental health among Somalis.

### Ethics

Ethical approval was granted by the King’s College London Research Ethics Committee and study recruitment ran from 05/06/2023 until 14/07/2023. An information sheet was provided to participants explaining the purpose of the study and written, informed consent was obtained. Three interviews were audio recorded. Four participants did not consent to being audio recorded so their interviews were recorded through note taking, including clarifying what participants had said by repeating this back to them. Pseudonyms were assigned to each participant to protect confidentiality.

## Results

This research aimed to gather better understanding of the perspectives of mental health among Somali people living in London. The open interview technique meant that a variety of questions were asked of participants, and different themes and opinions could be explored, with the participants expressing the significance of their own views. There were some themes that became evident within the interviews, including the personal importance of mental health to the participants, a concern around judgement in the community and the impact of shame and stigma on perceptions of mental health. When asked about the importance of mental health, all respondents expressed the significance of mental wellbeing in their lives, and reported opinions that robust mental health was as important as their physical health needs.

> It is very important because if you don’t take care of it, you have nothing left.

> When I compare my physical health to my mental health, I can’t function or do anything [when I’m mentally unwell].

One participant had lost a family member to a mental illness, whilst others described the stories of friends or family who had suffered with such conditions. Despite valuing mental health, three out of the seven participants were unable to list formal mental health conditions when asked, with two participants describing a need for a better education in mental health, either personally or within the community. For four participants, mental illness was discussed in regard to difficulty coping with life events, rather than any organic cause, with one participant saying, “when people choose that they can’t handle [stress], it goes to their mental [health]”. Furthermore, several participants differentiated between mental health conditions that they considered ‘severe’ and associated with madness, and those which they considered less severe, giving examples such as depression.

> [I] do acknowledge and understand the word mental health but do they mean depression, anxiety, low mood […] you could have a low mood and that could be your mental health.

> Yeah, there is no [such thing] as mental health, you know what I mean? Unless you are fully like a crazy person.

Furthermore, the use of the word ‘crazy’ and discussions of ‘madness’ was identified in the interviews. Some participants felt that this was a label the community would attach to them, if they spoke up about their mental health:

> You could be having like depression, or you can have like stress, or you don’t even have to have like full mental health yet and they will still label you as the crazy one.

Although this shows the concerns about stigmatisation from the community, it also demonstrates a distinction between “full” mental illness and conditions like stress. Prior research has found that Somali people tend to differentiate mental health into the ‘sane’ and ‘insane’, rather than perceiving mental health conditions as a spectrum [5]. This can stigmatise high-functioning individuals with mental health problems, since their condition does not affect their functioning within the community, and therefore could be seen as unimportant. Six participants used the words ‘crazy’ or ‘madness’ to discuss mental health conditions. This may result from a lack of Somali translation for some mental health conditions, but also could relate to this distinction of the ‘sane’ and ‘insane’. One participant implied that this vocabulary contributed to the stigma surrounding mental health, by linking public stigmatisation to concerns about individuals being able to fulfil their roles in society, saying:

> They’ll be like, she takes that [medication]. Why do you need that? Like she’s gonna go crazy. How do you know she’s not gonna kill your children when she gives birth?

The use of such labels contributes to the stigma surrounding mental illness, and perhaps poses the idea of mental health as a dichotomy between the sane and insane, rather than a spectrum involving a huge variety of conditions with different symptoms. Furthermore, this could lead to a difficulty engaging with mental illness diagnoses, for the fear of stigmatising labels such as ‘crazy’ being applied to the individual.

Indeed, stigma was a theme which predominated in the interviews, with all participants reporting a stigma within such discussions in the community. Six out of the seven participants used the word ‘shame’ in their responses, and all spoke at length about the opinions of community members on one’s mental health. Some participants described the effect of stigmatisation in isolating them from the community:

> They tend to ostracise a person that has [mental illness], sometimes they tend to make fun of you, and a lot of them they might say they are not practicing enough … prayers.

> You want to ask people for help and advice and first thing is you go to the community where you share the same values and norms […] But then it doesn’t really work out for the person because they are shamed.

The first response here directly references the ways that individuals are isolated from their communities, giving examples of them being ridiculed. Furthermore, this response explores the ways that community judgement on mental illness may involve religion, with concerns that speaking out about mental illness may cause judgement on the individual’s religious practices and that mental illness may simply be a result of not praying enough. Religion can be a helpful coping mechanism for those with mental illnesses, but views like this could also further isolate individuals with mental illness from religious communities. The second response reports the desire to seek help within the Somali community, demonstrating the importance of shared values to the individual, but remarking on the way that this could isolate an individual and undermine that desire for community connection.

Participants linked the idea of stigmatization to an intentional secrecy surrounding mental health conditions, with one saying:

> The family […] might tell you to hide it, don’t tell any other person because as a community if someone gets to know it, they will just talk bad about … our family.

This fear of judgement also affects the family’s reputation within the community. This may offer further reasons for individuals to feel concerned about discussing mental health with others and reinforce the taboo around mental health. Indeed, the theme of family was spoken about by all participants, with a range of responses noted. For four participants, family was considered important for supporting mental health recovery, with these participants reporting that they would approach their families if they were struggling. For the other three participants, however, family was not viewed as a support, with one participant saying, “Family can be worse” and another reporting that disclosing mental health conditions to family could cause an individual to fall out of favour. Given that research suggests that many Somali people will seek help from families and communities before mental health services [17], we should consider the impact that stigma may have on individual families and how it may affect their ability to care for a family member with a mental health condition.

In some interviews, comparisons were made to the treatment of mental health in Somalia, where some reported that individuals could be “chained to walls” or “hidden” in the family home. Three participants highlighted the idea of secrecy by discussing the practice of sending those with mental health conditions back to Somalia for treatment. In their reports, they described families telling the community that they had gone on holiday, gone to learn about their culture and religion, or gone away for work or study in order to hide their mental health issues.

> If someone gets help in the [UK] and people are shaming them in the way of getting married or going to work or to do anything, of course you are going to choose to go to another country to get the help, and then come back and hide it.

Importantly, this quote also references another theme from the interviews: that the stigma surrounding mental health could limit an individual’s opportunities, for example in work or marriage.

> If you’re not married, they will be like ‘oh, who wants to marry you, like you are crazy’. If you want to go to work, they will be like ‘who wants to hire you, you are crazy’ […] so people try to keep that to themselves.

Reports like that above demonstrate the effects of such stigma in limiting an individual’s ability to interact within their community, citing limitations to their roles in families or employment. With such a negative potential impact, this participant suggests that stigma can impact the course of a person’s life and contribute to the secrecy surrounding mental health conditions within the Somali community.

Participants suggested that there were differences between generations in their understanding and treatment of mental health conditions. Six participants reported instances of Somali elders denying the existence of mental health issues such as depression or stress. Some participants explained this in relation to older generations being raised in Somalia whilst younger generations were more likely to be raised in the UK. It was argued by three participants that older Somalis might reject Western interpretations of mental illness, particularly rejecting the medical model in favour of a religious or spiritual conceptualisation of mental illness.

> Still the older generation, they don’t believe stress […] they don’t believe mental illness, like they all, all they say is like go back to your God.

The interview data highlighted this generational difference, with participants attributing this difference to either older generations not wanting to know about mental health or being limited by the ways they were brought up. Indeed, one participant suggested that elders attributed mental illness to ‘westernised’ perspectives and lifestyles:

> You are too much in this modern world, that’s why you are like this.

> Did you get that from the European countries? Did you get that from the UK? Don’t bring that please because we don’t believe [it].

Here, the medical model of mental illness is presented as culturally inappropriate by older Somalis, which could contribute to stigmatisation of mental illness by creating a dichotomy between ‘local’ cultural tradition and ‘foreign’ Western medical model. Interview participants did not perceive these two perspectives as mutually exclusive, but complimentary, with three participants speaking about the importance of both medical treatment, such as medication and therapy, alongside cultural traditions, such as reading the Qur’an, praying and speaking to members of the community.

> Go to religion and the doctor and tell them – don’t hide anything.

Indeed, religion played a significant part in all interviews; all participants were Muslim and considered Islamic perspectives on mental health. For most respondents, this was connected to a religious duty to look after their mental wellbeing, with one participant saying, “Islam tells us to look after ourselves and tells us to get help if needed”. Other participants described the importance of prayer in supporting their mental wellbeing, with religion a huge source of comfort, and one participant suggested that religious scholars could help to break the taboo and stigma surrounding mental illness, by initiating discussions about mental health in the community. Spiritual concepts, such as Jinn or “black magic” were also mentioned in the interviews as being related to mental health. In contrast to the comfort of religious teaching, these concepts appeared to contribute towards the stigma of mental illness and were used to dismiss the validity of the concept. Furthermore, one participant spoke briefly about the concept of mental illness being seen a form of moral punishment, saying that others might ask:

> Why is their family like this? Maybe they did something wrong, so they deserve it.

One participant described her experience as a mother to a child with a mental health problem, and reported particular concern about younger generations, suggesting that they were more susceptible to mental health problems due to bullying and educational learning needs. On the other hand, many of the respondents spoke of the increased education concerning mental health among younger people, citing social media as both an educational service and a means of normalising mental illness within the Somali community. Furthermore, participants spoke positively about the provision of mental healthcare in the UK, the importance of free services such as therapy and the high number of Somali people working within healthcare. The participants thus spoke positively about the medical model of UK mental healthcare, and the importance of further understanding and education on the subject.

Results from these interviews identified a few key themes, with the most predominant being that of stigma and shame surrounding mental illness. Stigma was explored in relation to isolation from the community, intergenerational differences in conceptualising mental health and community and individual concerns around ‘westernised’ medical models of mental illness. In discussing mental illness, religion was seen as an important factor for encouraging mental wellbeing, ensuring support and helping fight stigma.

## Discussion

Stigma is a key element of Somali conceptualisations of mental health, and a significant barrier to individuals seeking formal psychological help [12,26]. Said et al. conceptualised stigma as either public or internalised, suggesting that public shame around mental illness leads to self-stigmatisation and secrecy concerning mental illness [26]. This was also seen in our research, where participants used the word ‘shame’ to describe community responses to mental illness, and the theme of secrecy – not revealing mental health struggles to others – was explored in six out of the seven interviews. Indeed, research has shown that community shame in mental health can impact the reputation of families and individuals by suggesting a relationship between weak faith and mental illness [26]. Similarly, in this research, one participant suggested that some community members might perceive mental illness as a result of weak faith, or “not practicing enough prayers”. It is important to note that this type of attribution within a religious community might also contribute to a sense of stigma or shame surrounding mental illness.

In our research, participants described the social impacts of stigma in limiting opportunities for employment and marriage. Michlig et al. similarly observed discussions in Somali focus groups which described people hiding their mental health problems to avoid perceived negative social consequences such as the threat of divorce [11]. Michlig’s research suggests that these perceived social consequences may lead to self-management of mental health conditions rather than accessing formal psychological help, whilst other research has also concluded that such attitudes can hinder help-seeking behaviours [18,26,27]. Moreover, other barriers to medicalised mental healthcare may come from a scepticism with seeking help outside of the community due to concerns that this might be considered shameful or culturally inappropriate [19,28]. However, participants in our research did not report problems with the UK’s medical model of mental health support, and all participants spoke positively of therapy, hospitals and medications, mentioning that these could help support mental wellbeing.

Alongside community preferences for community-based support, stigma could contribute to the practice of sending people with mental health conditions to Somalia to receive treatment. Research in Finland also found that the difference between medicalised models of mental illness in the Finnish healthcare system and Somali views of mental illness contributed to Somali people seeking mental healthcare abroad [29]. In our interviews, participants mentioned this practice, with some discussing the way that this might contribute to the secrecy surrounding mental illness. It has been observed that there is a cultural practice of sending individuals back to Somalia (sometimes referred to as *dhaqan ceelis*) if their behaviour is felt to be inappropriate, and concerns have been raised that this includes individuals with mental health conditions [30].

Differences have been observed in the Western understanding of mental health and the way the topic is understood ‘back home’ in Somalia, demonstrated by the denial of the existence of stress and depression in Somalia [16,26,31]. Despite recent improvements in the mental health training for Somali doctors, it is accepted that there is a lack of mental healthcare infrastructure in Somalia to support people with mental health conditions [32]. Thus, there is limited exposure to medical treatments, with traditional and religious healers acting as the main providers of mental healthcare [33]. Given this, Somali people who migrated later in life may bring support for practices learnt in Somalia with them to the UK. Indeed, research suggests that Somali people frequently keep traditional practices and health belief systems even after settling in a new country [2], which may explain the intergenerational differences in the understanding of mental health identified in our research.

These differing views may also contribute to the stigma surrounding mental health, with older generations conceptualising it as a product of Western culture [15]. Furthermore, problems of vocabulary may contribute to this generational divide, with an absence of words for mental health conditions in the Somali language, and reports of difficulty sharing such concepts even when interpreters are used [28]. Research on the elderly Somali population and their views of mental health are scarce, and more research would help to provide detail on this issue.

Many participants were able to convey the importance of religion to their understanding of mental health. There are established positive associations between mental health and religion, and prior research reports Somali people turning to faith, prayer and the Qur’an to help support their mental wellbeing [34]. In Somali regions of Africa, ilaajs – religious mental health facilities – are widespread and frequently the first choice of people seeking mental health services [35,36]. However, some participants did describe concerns around community responses to mental illness being seen as a distance from God, or moral weakness, a concern which has also been reported in prior research [16]. Although only one participant in this study brought up the concept of Jinn or possession, some researchers have reported concerns that spiritual conceptions of mental illness in Somali communities can further reinforce stigma [37].

It is important to note that prior research has found that family, traditional healers and religious leaders are normally primary caregivers for mental illness [19,36]. Given this, spiritual cures are often endorsed by Somali people, through prayer and reading of the Qur’an [2,26,27]. Furthermore, this community preference for religious and spiritual treatment may be helpful if integrated with medical treatments for mental health, and further research could be recommended in this area. Since participants may perceive scripture differently, further research may explore the connection between religious and cultural perceptions of mental health and the impact of religion on dismantling stigma and supporting mental health.

Stigma is an important barrier to mental health treatment that requires examination to allow for better community support. Research shows that family and community support can be protective against mental illnesses such as depression [38,39]. Specifically, research on East Africans in the United States found social support to be a key component to promoting psychosocial wellbeing, suggesting the detrimental effect of stigma and social isolation [40]. Social support has been posited as a means of ‘buffering’ stressful experiences, such as those, like trauma or discrimination, which migrants to the UK are more likely to have experienced [39,41]. In our interviews, participants discussed the effects of stigma on isolating them from the community, but most participants expressed a desire to have discussions about mental health within their families and communities, indicating the importance of dismantling stigma to create stronger community support.

## Conclusion

This research has identified some key themes surrounding perceptions of mental health among Somali people living in London. The main theme identified and reported by all participants was that of the shame and stigma surrounding mental health. On identifying this theme, further subthemes were observed, such as the impact of intergenerational differences, isolation from the community and the difficulties of integrating Somali and UK culture. Stigma around mental health was reported as both public and internalised shame and was consistent with prior research on Somali perceptions of mental health. Furthermore, participants discussed the impact of stigma in affecting their opportunities in employment and marriage and discussed the differences between the treatment of mental health in the UK and Somalia. Participants reported the importance of education on mental health within the community and suggested that religious and community leaders could help to dispel the stigma surrounding mental health. Religion was also discussed as a protective factor in mental wellbeing and promoted as a way of supporting those with mental health conditions. The significance of religion to Somali conceptions of mental healthcare is consistent with previous research and indicates the need for culturally appropriate mental health services, to help serve Somali communities in the UK.

Previous literature on the Somali community in the UK and their perceptions of mental health is somewhat limited, with much scope for further research. Opportunities for further research identified by this study include further studies on mental health stigma within Somali communities, research into intergenerational differences of opinion surrounding mental health, research identifying the mental health challenges for Somali adolescents and studies into the perception of different mental health treatments for Somali people. This is particularly important given the low service use in these communities and could offer further information on how best to organise mental health services to support and serve this community.

## Data Availability

Data will be held in public repository OSF.

## Acknowledgements

None

## Figures and Tables

Figure 1: location of the Somali diaspora. Pew Research Center (https://www.pewresearch.org/short-reads/2016/06/01/5-facts-about-the-global-somali-diaspora/). Accessed 16 February 2024.

## Data reporting

All data underlying the findings reported in this article will be shared through data repository OSF.

## Financial disclosure statement

This research was part of a student Master of Science programme and is unfunded.

## Notes

### Competing Interest Statement

The authors have declared no competing interest.

### Funding Statement

The author(s) received no specific funding for this work.

### Author Declarations

This study received ethical approval from King's College London Research Ethics Office, approval number MRSU-22/23-35179.

